# Assessing the impact of secondary school reopening strategies on within-school COVID-19 transmission and absences: a modelling study

**DOI:** 10.1101/2021.02.11.21251587

**Authors:** Trystan Leng, Edward M. Hill, Robin N. Thompson, Michael J. Tildesley, Matt J. Keeling, Louise Dyson

## Abstract

**Background:** Strategies involving rapid testing have been suggested as a way of reopening schools that minimises absences while controlling transmission. We assess the likely impact of rapid testing strategies using lateral flow tests (LFTs) on infections and absences in secondary schools, compared to a policy of isolating year group bubbles upon a pupil returning a positive polymerase chain reaction (PCR) test.

**Methods:** We developed an individual-based model of a secondary school formed of exclusive year group bubbles (five year groups, with 200 pupils per year). By simulating infections over the course of a seven-week half-term, we compared the impact of differing strategies on transmission, absences, and testing volume. We also considered the sensitivity of results to underlying model assumptions.

**Findings:** Repeated testing of year-group bubbles following case detection or regular mass-testing strategies result in a modest increase in infections compared to the policy of isolating year-group bubbles, but substantially reduce absences. When combined these two testing strategies can reduce infections to levels lower than would occur under year-group isolation, although such a policy requires a high volume of testing.

**Interpretation:** Our results highlight the conflict between the goals of minimising within-school transmission, minimising absences and minimising testing burden. While mass and targeted testing strategies can reduce school transmission and absences, it may lead to a large number of daily tests.

## 1 Introduction

To control the spread of SARS-CoV-2, the pathogen responsible for COVID-19, unprecedented restrictions have been placed upon people’s daily lives. In the UK, these non-pharmaceutical interventions (NPIs) have included practising good hand hygiene, wearing of face coverings, social distancing, the prohibition of households mixing socially, the restriction of a range of leisure activities, and the closure of educational establishments, workplaces, pubs and restaurants. Collectively, such measures have reversed the growth in infection during the first and subsequent epidemic waves.

While these measures, together, can be effective at controlling the spread of infectious diseases, their implementation has come with significant societal and economic costs^1,2^. In particular, the closure of schools puts educational outcomes at risk, especially for disadvantaged pupils, with existing inequalities and attainment gaps being exacerbated^3^. School closures have a particularly adverse impact on vulnerable children due to reduced access to essential services^4^, and impair the physical and mental health of many children^5^. The closure of schools has also placed an additional strain on parents, particularly for single-parent households, and the burden of care during lockdowns has exacerbated gender inequities^6^. Owing to these harms, the UK government has expressed that keeping schools open is a priority in their plans.

Given the societal benefits of keeping schools open, a variety of strategies aiming to minimise transmission within the school setting have been considered. In the UK, a range of measures have been introduced to schools^7^, including socially distanced spacing between desks, restricting the movement of pupils within the school, restricting social mixing between year groups, mandatory mask wearing in thoroughfares, as well as an increased emphasis on hand washing and general hygiene measures. From September to December 2020, to halt chains of transmission within a school, upon a pupil receiving a positive polymerase chain reaction (PCR) test, other pupils who have been in close contact with the positive case have had to isolate for 10 days (14 days originally). These ‘close contacts’ have typically referred to an entire year group bubble, although some schools have implemented more targeted approaches^8^. While these measures are expected to have been effective in reducing transmission, they have also led to a considerable number of school days missed throughout the term. These absences have an impact on childrens’ education, the ability to assess pupils fairly, and undermine the benefits of keeping schools open.

For staff and pupils returning to secondary schools in England during January 2021, it was planned for rapid coronavirus testing to be introduced using Lateral Flow Device tests (LFTs). It was originally envisaged testing would be provided (as a pair of LFTs) for all secondary pupils and staff prior to a return to face-to-face teaching. After this, staff would be tested once a week on an ongoing basis. Additionally, should a pupil receive a positive test, all close contacts of the pupil would be tested using an LFT for the next seven days. Positive tests identified during this period would trigger a further round of testing, until no new cases were been identified for a period of seven days^9^. By identifying and isolating asymptomatic and presymptomatic individuals, it is hoped that a mass testing strategy would be effective in controlling transmission within schools while minimising absences. Additionally, doing so may identify a large proportion of infected individuals, helping to facilitate an improved test and trace strategy.

Mass testing strategies have been implemented in other contexts, with mixed success. Preliminary data released from the field evaluation of testing in asymptomatic people in the city of Liverpool indicated just 48.89% of COVID-19 infections in asymptomatic people were detected with a Innova Lateral Flow SARS-CoV-2 antigen test, when compared with a PCR test^10^. Furthermore, the antigen test failed to detect three in ten cases with the highest viral loads. Testing has also been a constituent part of guidance for the movement of university students to and from universities^11^. The guidance stipulated all students should be offered rapid tests before leaving and when they return to university to help identify and isolate those who are asymptomatic but could spread the virus. The protocol involved two LFTs, three days apart.

The role of children in the spread of COVID-19, particularly in the school setting, remains unclear. Evidence from a range of sources suggests that children are, in general, only mildly affected by the disease and have low mortality rates^12^. This is reflected in the fact that by 27th January 2020 there had been 69, 801 people who had died in hospitals in England and had tested positive for COVID-19, but only 29 of those were in the 0-19 year age group^13^. Additionally, SARS-CoV-2 infections and outbreaks were uncommon in educational settings during the summer half-term in England^14^, with the available evidence suggesting that transmission among children in schools can happen but less efficiently than other respiratory viruses such as influenza^15^. Similar signals have been noted in other nations. However, a synthesis of the literature by the European Centre for Disease Prevention and Control attributed moderate confidence to onward transmission by adolescents occurring as often as by adults in household and community settings, given social mixing patterns; they also indicated, though with weaker support, that preschool and primary school aged children transmit SARS-CoV-2 less often than adolescents and adults^16^. There are complexities with balancing harms associated with school openings resulting in increased transmission, with harms associated with closure of educational establishments stunting the educational development and welfare of children.

To assess the potential impact of a strategies involving rapid testing on within-school transmission and absences, we created an individual-based model of a secondary school formed of exclusive ‘year group bubbles’. By performing computational simulations corresponding to infection spread over the course of a school half-term, we compared the impact of strategies involving rapid testing to a strategy of isolating year group bubbles on the total number of infections within schools, the number of school days missed per pupil and the proportion of infected individuals identified. By doing so, we offer an assessment of the relative merits of a mass testing strategy compared to an isolation of year groups strategy. We also considered the sensitivity of our results to underlying model assumptions. Consequently, we identify factors likely to have the largest impact on the success of a mass testing strategy.

## 2 Methods

In this study, we used a discrete-time stochastic individual-based model, with a daily time step, to simulate the spread of infection within a secondary school over a half-term of seven weeks. In our simulations, schools consisted of five year groups, with each year group containing 200 pupils, equivalent to a secondary school without a sixth form (ages 11-16, the inclusion of additional year groups representing a sixth form does not qualitatively change results). We provide a summary of our baseline assumptions, as well as the sensitivity assumptions we considered, in Table 1.

**Table 1:** **Description of model parameters, our baseline parameterisation and the alternative values considered in the univariate sensitivity analysis**.

| Description | Baseline | Sensitivity | Source |
| --- | --- | --- | --- |
| LFT sensitivity | 30 day positive test probability profile for symptomatics<br>Faster decay in positive test probabilities for asymptomatics (Supporting Text S2) | 95% credible intervals from<sup>25</sup> for symptomatics<br>Transformed 95% credible intervals from<sup>25</sup> for asymptomatics | 25,28 |
| PCR sensitivity | As above | As above | 25,28 |
| within-school transmission, $K$ | Unif(1,4) | Baseline - $K = 2.5$<br>Low - $K = 1$<br>High - $K = 4$ | Assumption |
| External source of infection, $\epsilon$ | 0.00131<br>(such that 10% of individuals are infected over a half term) | Low - $0.5 \times \epsilon$<br>High - $2 \times \epsilon$ | Assumption |
| Relative probability of transmission since day of infection | $\Gamma(5.62, 0.98)$ | - | 17 |
| Incubation period (time until symptom onset) | $\Gamma(5.807, 0.948)$ | - | 20 |
| % of pupils symptomatic | Unif(12,31) | Baseline - 21.5%<br>Low - 12%<br>High - 31% | 18 |
| Relative infectiousness of asymptomatic individuals (%) | Unif(30, 70) | Baseline - 50%<br>Low - 30%<br>High - 70% | 19 |
| Initial population level immunity | 20% | Low - 10%<br>High - 30% | 23 |
| Initial population level prevalence | 2% | - | 22 |
| Interaction between year groups, $\alpha$ | 0 - i.e. 100% of within-school infections are within-year | 1 - i.e. 20% of within-school infections are within-year | Assumption |

We assumed that simulated schools implemented a ‘bubbling’ policy at the level of year groups. In our baseline scenario, we assumed exclusive and effective year group bubbles, meaning that there was no transmission between year groups, but pupils mix randomly within year group bubbles. We did not explicitly model teachers, siblings, or external contacts. The impact of teachers and siblings can be indirectly captured by assuming some degree of transmission between year groups, a scenario considered in our sensitivity analysis. Infected pupils’ relative probability of within-school transmission since day of infection was derived from data from known source-recipient pairs^17^, with an assumed incubation period distribution under the assumption that the generation time and incubation period are independent. The level of onward transmission within-school remains unclear, and is likely to be influenced by a variety of factors, including the success of other within-school social distancing measures and the epidemiological characteristics of the dominant strain of SARS-CoV-2 in circulation in the local area. Because of this, we considered a wide range of levels of within-school transmission. We assumed that the impact of external contacts on transmission could be captured by a constant external force of infection, chosen to satisfy an average of 10% of pupils becoming infected by the end of the half-term under an isolation of year groups policy. When isolating, we assumed that individuals adhered and effectively isolated. After 15 days, we assumed individuals were no longer infectious and recover with immunity. For a more detailed summary of our model’s assumptions regarding transmission, see Supporting Text S1.

Informed by previous studies into the levels of asymptomatic infection within age-groups, we assumed that 12-31% develop symptoms over the course of their infection^18^, with the rest of the school population remaining asymptomatic. There are indications that asymptomatic individuals may be less infectious than symptomatic individuals^19^. Accordingly, we assumed that asymptomatic pupils were 30-70% as infectious as those that develop symptoms. Symptomatic pupils developed symptoms on a day drawn from a Gamma distribution with shape 5.807 and scale 0.948^20^, corresponding to a mean time to symptom onset of 5.5 days. We assumed that the relative probability of transmission of an individual and the time to symptom onset were independent, though in reality these factors likely influence one another^21^. Under our assumptions, approximately 50% of infectiousness occurred during an individual’s presymptomatic infection phase. In line with recent observations from community surveillance surveys, we assumed that the population prevalence at the start of the simulation was 2%^22^ and that 20% of the population had been previously infected and as a result were immune from reinfection (based on an estimate from December 2020 that approximately 12% of the population in England would have tested positive for antibodies to SARS-CoV-2 from a blood sample^23^, with an expectation the true proportion previously infected would be higher owing to waning of detectable antibodies^24^).

Upon symptom onset, infected pupils underwent a PCR test. Pupils self-isolated until they received a test result, and we assumed that pupils received a result two days after taking a test. Those receiving a negative result returned to school the day after receiving their result, while those testing positive entered isolation for ten days. Pupils who tested positive using a LFT entered isolation, with the outcome of a confirmatory PCR test then determining whether the pupil remained in isolation or was released from isolation. Accordingly, we assumed that identified infected pupils did not transmit infection on the day they were tested. We used previously estimated LFT and PCR test probability profiles for symptomatic individuals^25^. For asymptomatic individuals, we assumed that the probability of testing positive is equal to that of symptomatic individuals until peak positive test probability, but then decays more rapidly (Supporting Text S2). We assumed a PCR test specificity of 100%, in line with recent studies confirming that false positives from PCR are rare^26^). We assumed LFT specificity to be 99.7%^27^.

Using this model, we assessed the impact of different reopening strategies on transmission and absences within schools. By simulating different school return strategies in simulations with the same set of parameter values and same set of pregenerated random numbers, we could directly compare the impact each strategy had for each simulation. Specifically, we produced 10,000 simulation replicates for five strategies: (i) isolation of year group bubbles; (ii) serial contact testing; (iii) regular mass testing; (iv) a combination of regular mass testing with serial contact testing; (v) no school-level testing or isolation of year-group bubbles. Under an isolation of year group bubbles strategy, upon the identification of a case (through a symptomatic pupil seeking a PCR test), all pupils within a year-group bubble were placed in isolation for ten days following the last contact with the positive case. For serial contact testing, on the identification of a case (following confirmation by PCR), all pupils within a year-group bubble were tested using LFT for seven days following the last contact with the positive case. The period of serial contact testing is reset if another pupil in the year-group bubble returned a positive LFT result. Under a regular mass testing strategy, all pupils are tested once a week using LFTs. For strategies (ii), (iii), and (iv), all pupils were tested twice with LFTs in the week before returning to school. We elaborate on the details of each reopening strategy in Supporting Text S3.

In addition, we assessed the sensitivity of results obtained to the underlying modelling assumptions (see Supporting Text S4) and we also explored the impact of altering the assumed level of within-school transmission (Supporting Text S5). For the sensitivity analysis, we generated 2000 simulation replicates for each scenario. We performed the reopening strategy model simulations, sensitivity analysis and visualisation of results using Matlab 2019b.

## 3 Results

In the majority of the 10,000 simulations (85.8%), a serial contact testing strategy was less effective at reducing infections within a school than an isolation strategy. Similarly, in 89.8% of simulations, regular weekly testing was less effective at reducing infections than isolating year group bubbles. However, serial contact testing and regular testing combined was more effective at reducing infections than isolating year group bubbles in 92.5% of simulations (Figure 1a).

**Figure 1:**
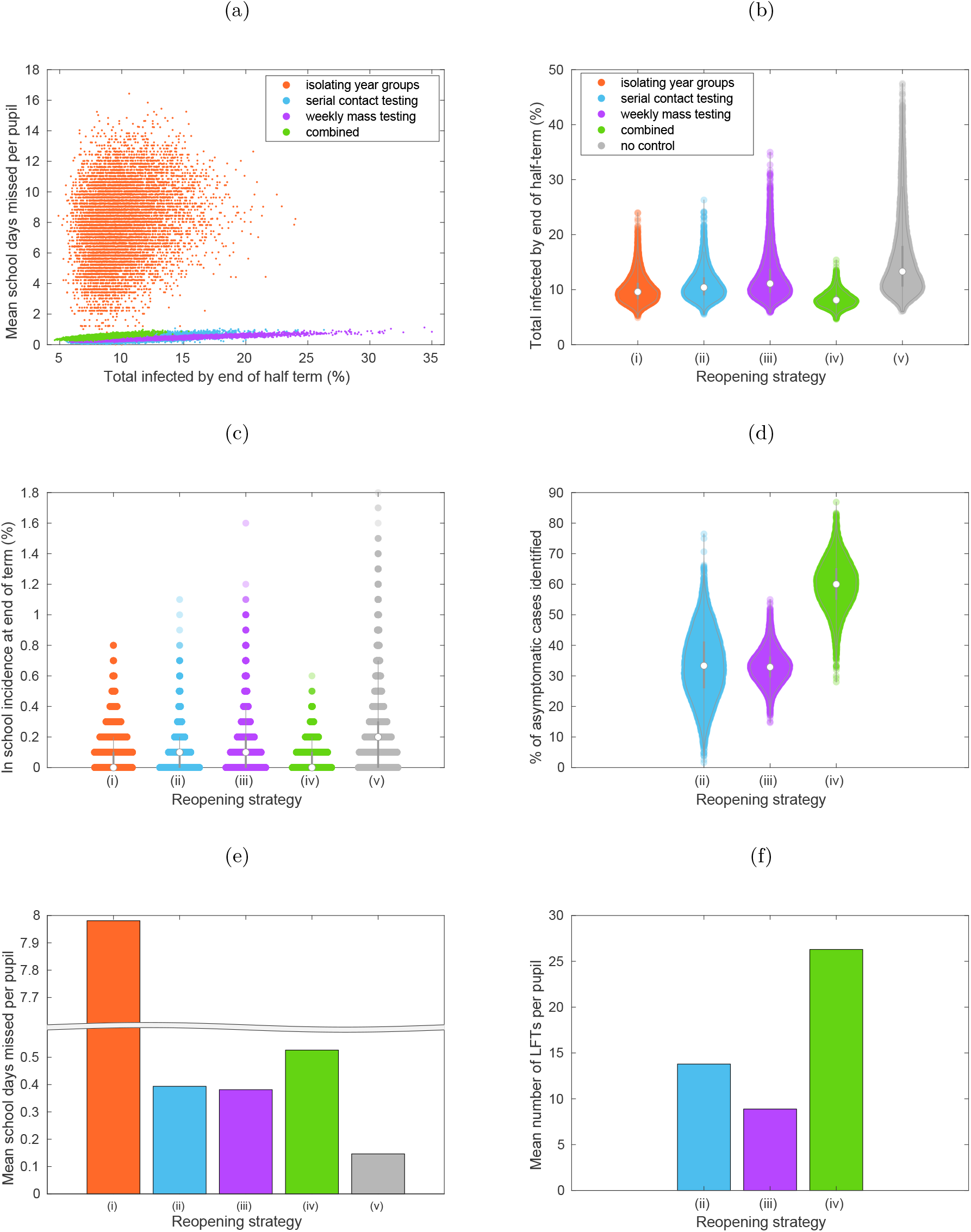
The trade-off between transmission, absences, and testing volume. **(a)** Relationship between total infections and school days missed for an isolation of year group bubbles strategy (orange), serial contact testing (blue), weekly mass testing (purple), combined serial contact testing and mass testing (green). Strategies including rapid testing minimise the average number of school days missed per pupil, yet also correspond to a larger number of total infections. **(b)** Violin plots of percentage of school pupils infected during the course of the half-term.**(c)** Violin plots of within-school incidence at the end of the half-term. **(d)** For strategies involving rapid testing, violin plots of the percentage of asymptomatic cases that had been identified through rapid testing by the end of the half term. **(e)** The mean number of school days missed per pupil. **(f)** For strategies involving rapid testing, the mean number of LFTs taken per pupil. Results were obtained from 10,000 simulations.

The average number of pupils infected over by the end of the half-term for an isolation of year-group bubbles strategy was set at 10%. Slightly higher levels resulted from a serial contact testing (10.82%) or regular weekly testing strategy (12.86%), whereas lower levels resulted from a combination of both measures (8.20%). Without control measures, a mean of 15.21% had been infected by the end of the half-term, with large outbreaks occurring much more frequently (Figure 1b). These trends are reflected by the within-school incidence at the end of the half-term; with similar within-school incidence for isolation and serial contact strategies, reduced within-school incidence for a combined strategy, and substantially higher within-school incidence with no control strategy (Figure 1c).

By identifying asymptomatic and presymptomatic individuals, mass testing prior to the start of term initially reduced mean prevalence within schools. However, prevalence quickly increased to a higher level than would be obtained under an isolation of year group bubbles strategy, remaining at a higher level for weekly mass testing and serial contact testing strategies from the middle of week 2 onwards (Figure 2a). Under both strategies, a considerable proportion of asymptomatic infected individuals remained unidentified throughout the simulation (Figure 1d, Figure 2b). By combining serial contact testing and regular mass testing, the majority of asymptomatic individuals were identified over the course of the half-term, and prevalence remained low throughout the course of the half-term.

**Figure 2:**
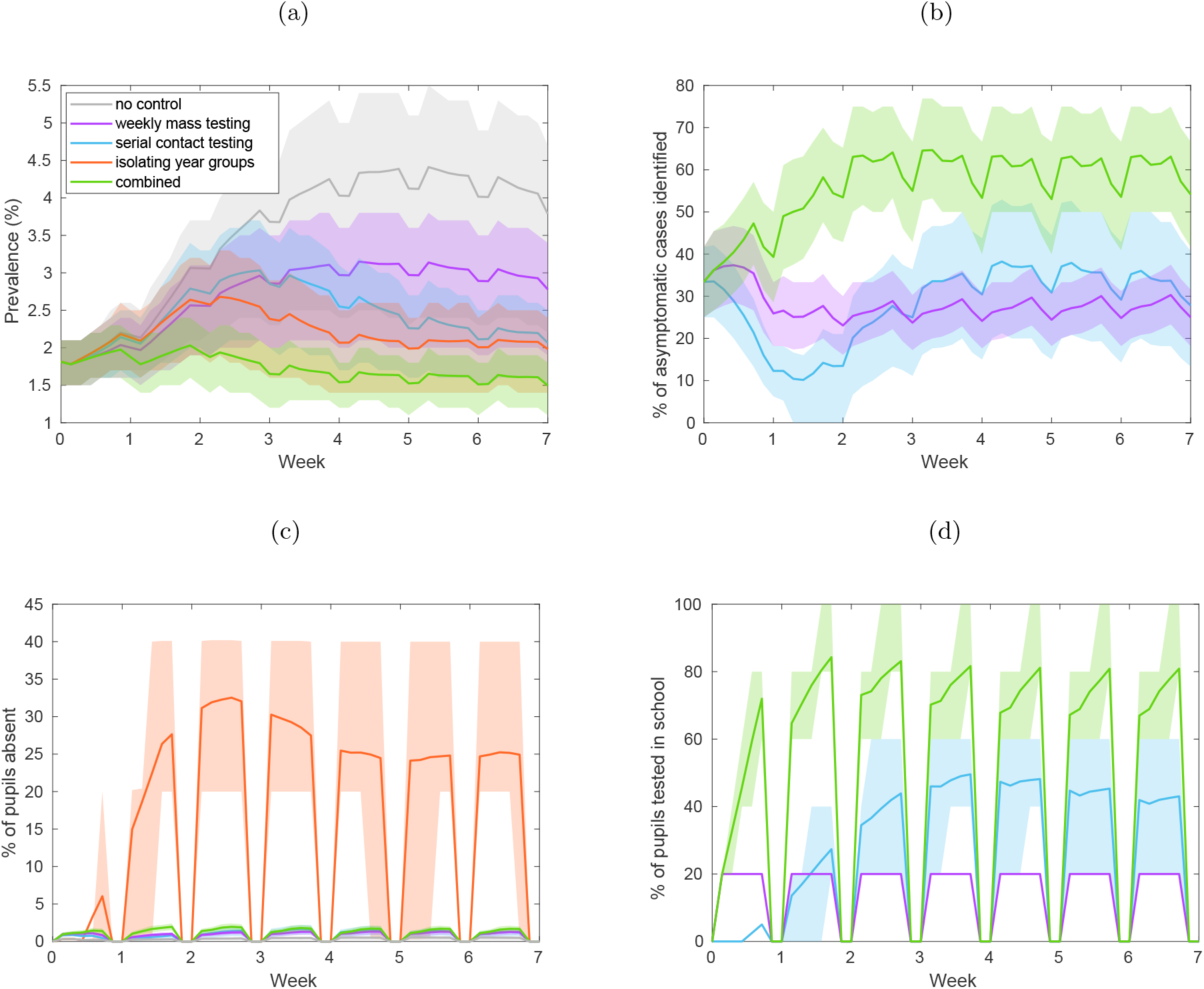
Infection, absences, and testing over the duration of the school half-term. We display timeseries of **(a)** prevalence, **(b)** the percentage of currently infectious asymptomatic individuals identified for reopening strategies involving within-school testing. We display timeseries of **(c)** the percentage of pupils absent and **(d)** percentage of pupils tested throughout the half-term. Solid line traces correspond to the mean value attained on each daily timestep and shaded envelopes represent the 50% prediction intervals (these regions contain 50% of all simulations at each timepoint). The strategies displayed are: no control (grey), weekly mass testing (purple), serial contact testing (blue), year group bubbles strategy (orange), combined serial contact testing and mass testing (green). Results were obtained from 10,000 simulations.

Serial contact tracing and mass testing strategies were more effective at reducing school absences than an isolation of year group bubbles approach. By isolating year group bubbles, even uninfected pupils can spend a considerable number of days absent. The average pupil spent 7.98 days isolating over the duration of the half term, i.e. around 22.8% of a half-term of seven weeks. For serial contact tracing and mass testing strategies, as individuals would only be absent if they had sought a PCR test, or had tested positive to an LFT or PCR test, the majority (91.9% under serial contact testing, 92.8% under weekly mass testing, 88.0% under a combined strategy) of pupils had no days of school absence. The mean days absent for these strategies was 0.39 days for a serial contact testing strategy, 0.38 days for regular weekly testing, and 0.53 days for those measures combined (Figure 1e). Temporally, for a isolation of year group bubble strategy, throughout the half-term a considerable portion of pupils (20-40%) may plausibly be expected to be absent. For the considered strategies involving rapid testing, the fraction of students absent at any one time was relatively low, remaining below 5% throughout the half-term (Figure 2c).

Strategies involving rapid testing required a large number of tests to be implemented successfully. Over the course of a half term, pupils on average undertook 13.8 LFTs under a serial contact testing strategy, 8.9 LFTs under a weekly mass testing strategy, and 26.3 LFTs under a combined strategy (Figure 1f). For strategies involving serial contact tracing, a high volume of testing was required throughout the half-term (Figure 2d). This high volume of testing also required many pupils to isolate over weekends, when tests could not be administered.

There is considerable uncertainty surrounding many of the parametric assumptions that underpin the model. Accordingly, we performed a univariate sensitivity analysis to understand the impact these assumptions have on our findings (Supporting Text S4). Across the range of alternative parameterisations considered, serial contact testing and regular mass testing strategies remained less effective at reducing infections than an isolation of year group bubbles strategy alone, but remained more effective when combined. Of all the factors considered, the sensitivity of LFTs had the largest impact on infections. Community infections, captured by the external force of infection on individuals, and the level of within-school infection, had the largest impact on the reduction of school days missed. We also explored the impact of altering the assumed level of within-school transmission (Supporting Text S5). Different levels of within-school transmission did not qualitatively change findings, but identifying asymptomatic infectious individuals had a larger benefit for higher quantities of within-school transmission.

## 4 Conclusions

In this paper, we have developed an individual-based model of a secondary school formed of exclusive ‘year group bubbles’ and performed numerical simulations to assess the impact of a collection of postulated testing and isolation-based school control strategies (against spread of SARS-CoV-2) on transmission, absences and testing burden. Across the considered strategies, our findings reveal a trade-off between these three measures.

Evaluating strategies on the basis of school absence, serial contact testing and regular mass testing reduce absences considerably. However, these approaches adopted individually result in slightly higher levels of infection than would be obtained under an isolation of year group bubbles strategy. Used together these two test-based strategies can result in lower levels of infections but both strategies, particularly when implemented in tandem, require a high testing capacity. Prior work performing numerical simulations on complex networks has indicated a high volume of testing being required to effectively curb disease spread^29^.

In comparison to isolating year group bubbles breaks chains of transmission when a positive case is identified, rapid testing strategies can allow infected pupils who falsely test negative to continue to transmit infection within the school setting. Under serial contact testing, asymptomatic individuals may fail to be identified for two reasons. Firstly, the sensitivity of LFTs means that some asymptomatic individuals would remain undetected despite being tested repeatedly while infected^10^. Secondly, serial contact testing is only triggered after a positive case is identified by other means. As a substantial proportion of the school population will remain asymptomatic throughout their infection, there is the possibility that infection continues within the school setting for a considerable period before an individual is infected who exhibits symptoms. In contrast, testing before term starts, and then regularly testing throughout the half-term, reduces the risk of large chains of transmission occurring unnoticed. By combining this with initiating serial contact testing, enough asymptomatic and presymptomatic individuals are identified to keep incidence within the school low.

We have underpinned our model with relevant available data where possible and performed a sensitivity analysis to understand the sensitivity of model outputs to the underlying model assumptions. Our results did not qualitatively change across the range of sensitivity assumptions considered. Of all the factors considered in our sensitivity analysis, the sensitivity of LFTs had the largest impact on infections. While previous modelling approaches have shown that the frequency of test screening has a larger impact on reducing transmission than test sensitivity^30^, our results demonstrate that the sensitivity of rapid tests may still be an important determinant of the relative effectiveness of different school reopening strategies that involve rapid testing at reducing transmission compared to strategies involving isolation. We believe these outcomes are reconciled by the differing model approaches affecting the relative impact of test sensitivity; the study by Larremore *et al*.^30^ assumed that test results were a deterministic function of viral load, while in our model the likelihood of an individual testing positive was governed by a probability distribution dependent on the time since infection (inferred from observed data from UK healthcare workers^25^). These alternative approaches result in different levels of infectiousness removed through rapid testing, as under the assumptions of Larremore *et al*. individuals with high viral loads will always test positive to LFTs. This highlights the importance of continued research into the sensitivity of rapid tests, with granularity to determine heterogeneity (if any) across specific age groups and in specific settings, as well as the most appropriate way to capture test probability profiles in a model.

Our model makes several optimistic assumptions regarding the practicalities of testing. Schools are assumed able to test pupils in an infection-secure environment, and able to effectively isolate positively identified pupils from passing on infection within the school on the day of testing. We assume that the sensitivity of LFTs that are self applied by pupils is comparable to that of LFTs self applied by healthcare workers^25^. In our model, pupils required to isolate do so effectively, and have no risk of becoming infected while isolating. All pupils comply with the school’s reopening strategy, and all symptomatic pupils seek a PCR test and isolate upon symptom onset. By doing so, we demonstrate the impact of reopening strategies if ideally implemented. However, in reality this ideal scenario is unlikely to be met. If pupils have a substantial chance of transmitting infection to other pupils when being tested in school, or do not take swabs correctly, this will increase the infections that occur when implementing a serial contact testing or regular mass testing strategy. Conversely, if pupils have a substantial risk of infection when they are supposed to be isolating, a strategy that keeps pupils within school, where social interactions are regulated, may become more beneficial. For strategies involving serial contact testing, pupils are expected to isolate on days that they are due to be tested that fall on weekends. Pupils failing to isolate on serial contact test days that fall on weekends will reduce the effectiveness of these approaches. The impact of other forms of non-adherence will likely depend upon schools’ policies regarding non-adhering pupils.

Whilst this study has focused on the impact of reopening strategies in secondary schools, results may be expected to be qualitatively similar in the context of primary schools. However, there are some key differences that may impact the appropriateness of applying our results directly to a primary school setting. As tests within a school are expected to be self-administered, this may not be feasible for primary school age children, particularly in younger years. Epidemiological and clinical factors may differ between primary and secondary school aged children^16^, meaning the effectiveness of serial contact testing, which often relies upon testing being initiated by a symptomatic case seeking treatment and testing positive, may be affected. The relevant size of the exclusive bubbles in primary schools will typically be smaller, as pupils are often partitioned into exclusive bubbles based on individual classes rather than entire year groups. Use of smaller exclusive bubbles could potentially impact both transmission and testing.

An evaluation of the risk of reopening strategies to teachers and the wider community is beyond the remit of this study, though this risk will be a function of the level of infections that result from different reopening strategies. Capturing this aspect of transmission would require the explicit modelling of teachers and external contacts of both pupils and teachers. Research into these risks would be a valuable line of enquiry for future research, but as is often the case, the challenge would be the appropriate parameterisation of the model.

In summary, we have explored the impact of different reopening strategies on both transmission and absences within a secondary school setting. We find that serial contact testing and regular mass testing strategies, acting alone, are less effective at reducing infections than an isolation of year group bubbles strategy, but substantially reduce absences. Acting together, serial contact testing and regular mass testing can reduce infections to levels lower than would occur under an isolation of year group bubbles strategy, but such a policy requires a high volume of testing. Our results highlight the conflict between the goals of minimising within-school transmission, minimising absences, and minimising testing burden. Amongst the strategies considered here we did not identify one that minimised all three simultaneously. An assessment of the relative benefits and costs of each must be made when considering future school reopening policies.

## Supporting information

Supporting information for the main text

## Data Availability

Data used to parameterise the study is publicly available and stated within the main manuscript and Sup- 295
porting Information. Code for the study is available at: https://github.com/tsleng93/SchoolReopeningStrategies

## Author contributions

**Conceptualisation:** Trystan Leng; Edward M. Hill; Robin N. Thompson; Michael J. Tildesley; Matt J. Keeling; Louise Dyson.

**Data curation:** Trystan Leng; Edward M. Hill.

**Formal analysis:** Trystan Leng.

**Funding acquisition:** Matt J. Keeling; Michael J. Tildesley; Louise Dyson.

**Investigation:** Trystan Leng.

**Methodology:** Trystan Leng; Edward M. Hill; Robin N. Thompson; Michael J. Tildesley; Matt J. Keeling; Louise Dyson.

**Software:** Trystan Leng; Edward M. Hill.

**Supervision:** Michael J. Tildesley; Matt J. Keeling; Louise Dyson.

**Validation:** Trystan Leng; Edward M. Hill.

**Visualisation:** Trystan Leng.

**Writing - original draft:** Trystan Leng; Edward M. Hill.

**Writing - review & editing:** Trystan Leng; Edward M. Hill; Robin N. Thompson; Michael J. Tildesley; Matt J. Keeling; Louise Dyson.

## Data Availability

Data used to parameterise the study is publicly available and stated within the main manuscript and Supporting Information. Code for the study is available at: https://github.com/tsleng93/SchoolReopeningStrategi

## Supporting information items

### Supporting Text S1

**Transmission dynamics in detail**. Description of infectiousness over time, the parameterisation of within-school transmission, external infection, interaction between year groups, and recovery and immunity.

### Supporting Text S2

**Test probability profiles for symptomatic and asymptomatic individuals**. LFT and PCR test probability profiles and the methods used to obtain asymptomatic test probability profiles.

### Supporting Text S3

**Reopening strategies in detail**. A description of each reopening strategy considered.

### Supporting Text S4

**Sensitivity Analysis**. Description of the univariate sensitivity analysis performed, and the results from this analysis.

### Supporting Text S5

**Level of within-school transmission**. The impact of different levels of within-school transmission, while keeping the total number of infections under an isolation of year group bubbles strategy constant, on different reopening strategies is explored.

