## Supporting information for the main text for "Assessing the impact of secondary school reopening strategies on within-school COVID-19 transmission and absences: a modelling study"

#### Table of Contents

**1 Supporting Text S1: Transmission dynamics in detail** **2**

**2 Supporting Text S2: Test probability profiles for symptomatic and asymptomatic individuals** **4**

**3 Supporting Text S3: Reopening strategies in detail** **5**

**4 Supporting Text S4: Sensitivity analyses** **6**

**5 Supporting Text S5: Level of within-school transmission** **10**

### 1 Supporting Text S1: Transmission dynamics in detail

#### Infectiousness over time

In our model, infected pupils attending school (i.e. those not isolating and it being a school day) transmit infection to other pupils within their year group with a probability dependent on the time elapsed since their infection. Specifically, we assume that the relative probability of transmission since the day of infection is given by a Gamma distribution with shape 5.62 and scale 0.98. This distribution was derived from data from known source-recipient pairs [1], with an assumed incubation period distribution (Gamma distributed with shape 5.807 and scale 0.948 [2]) under the assumption that the generation time and incubation period are independent.

#### Parameterising within-school transmission

We defined the level of transmission within a school by a parameter, $K$. Specifically, $K$ defined the expected number of secondary cases from an infected symptomatic pupil, assuming a fully susceptible school population and that the impact of depletion of susceptibles is negligible; in reality, due to the small size of school populations, the depletion of susceptible individuals is never negligible, and hence an infected symptomatic pupil would be expected to infect less than $K$ other pupils. The appropriate choice of $K$ is unclear, and is likely to be influenced by a variety of factors that may vary from school to school, including the success of other within-school social distancing measures and the epidemiological characteristics of the dominant strain of SARS-CoV-2 in circulation in the local area. Due to this uncertainty, we considered a range of different values of $K$, specifically, $K \sim Unif(1, 4)$. We assumed that asymptomatic individuals are between 30 and 70% as infectious as those that develop symptoms, i.e. if a symptomatic pupil is expected to infect $K$ other pupils over the course of their infection, an asymptomatic pupil is expected to infect $0.3 - 0.7 \times K$ other pupils.

#### External infection

As well as transmission within school, we assumed that all pupils who were not isolating had a constant probability of external infection each time step, denoted $\epsilon$, representing the force of infection to pupils from the wider community. In our baseline parameterisation, we set $\epsilon$ such that an average of 10% of pupils became infected by the end of the half-term under an isolation of year groups policy. When isolating, we assumed that individuals adhered and effectively isolated, meaning they had no probability of becoming infected whilst isolating.

#### Interaction between year groups

The impact of interaction between year groups is captured by the parameter $\alpha$ in our model. We let $I_j(t, k)$ denote the probability that an individual $k$ in year group $j$ on day $t$ infects another pupil (assuming that all other pupils are susceptible), which will depend on the day individual $k$ was infected, whether they are symptomatic or asymptomatic, whether they are isolating, and whether it is the weekend. Assuming the exclusivity of year-group bubbles (our baseline assumption), and assuming frequency-dependent transmission, the probability of infection to any given individual in year group 1, which we denote $\beta_1$, is then given by:

$$\beta_1(t) = \frac{\sum_k I_1(t, k)}{N_1} \tag{1}$$

On the other hand, assuming that there is random mixing between year groups, $\beta_1$ satisfies:

$$\beta_1(t) = \frac{\sum_i \sum_k I_i(t, k)}{\sum_i N_i} \tag{2}$$

Collectively, we define the parameter $\alpha$ as follows:

$$\beta_1(t) = \frac{(\sum_k I_1(t,k) + \alpha \sum_{i=2}^{5} \sum_k I_i(t,k))}{N_1 + \alpha \sum_{i=2}^{5} N_i} \tag{3}$$

such that for $\alpha = 0$, Eq. (1) is satisfied, and for $\alpha = 1$, Eq. (2) is satisfied.

#### Recovery and immunity

After 15 days, we assumed individuals were no longer infectious and recover with immunity. Whilst the duration of immunity conferred from SARS-CoV-2 infection remains uncertain, infection appears to (on average) confer a period of immunity at least of the order of months long[3].

#### 2 Supporting Text S2: Test probability profiles for symptomatic and asymptomatic individuals

While the probability of testing positive through time has been detailed for both PCR and LFT tests, prior analyses have typically been based on symptomatic individuals[4]. The probability of testing positive is likely a function of viral load; while symptomatic and asymptomatic individuals have similar average peak viral loads and proliferation stage durations, their average duration of clearance stages has been observed to differ[5,6]. In this paper, we assumed that the probability of asymptomatic individuals testing positive to both PCR and LFT was equal to that of symptomatic individuals until the peak of infection, but then decays more rapidly, such that the probability of an asymptomatic individual testing positive at 6.7 days after the peak should equal the probability of a symptomatic individual testing positive at 10.5 days after the peak (corresponding with estimates that the average duration of clearance of 10.5 days is symptomatic cases and 6.7 days in asymptomatic cases[5]). We applied the same method to the 95% credible intervals provided by[4] to obtain high and low sensitivity test probability profiles. However, we express that this is an area of considerable uncertainty. Future studies detailing the testing probability of asymptomatic individuals, and the specific relationship between viral load and testing probability, would be a valuable contribution to this area.

As a final step, we adjusted for the specificity of LFTs by setting the minimum daily probability of testing positive to be 0.3%[7].

**(a)** **(b)**

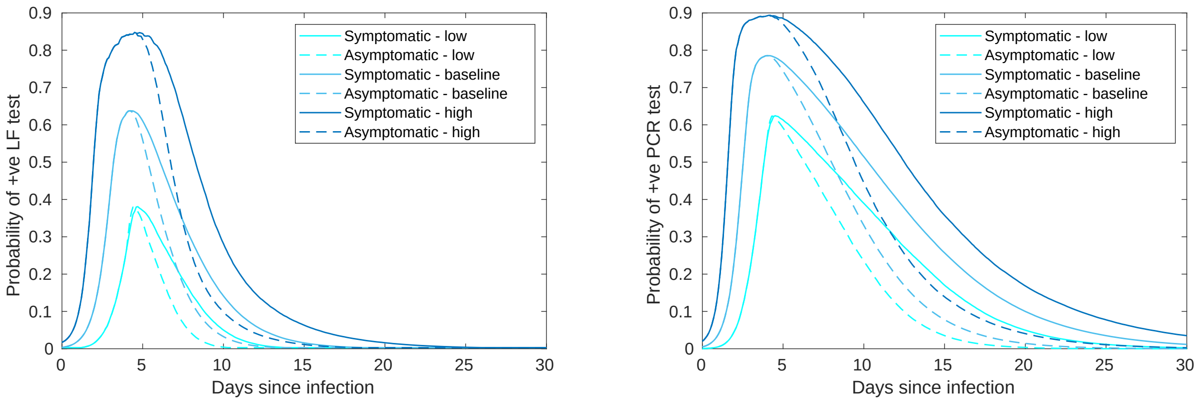


**Fig. S1: Probabilities of testing positive through time for symptomatic and asymptomatic individuals.** We assumed that the probability of positive **(a)** LFTs, and **(b)** PCR tests in symptomatic and asymptomatic individuals were equal during the proliferation stage of the virus, but that the probability of asymptomatic individuals testing positive decayed faster in the clearance stage, owing to a shorter mean clearance duration of 6.7 days[5]. We took the above profiles for symptomatic individuals directly from[4]

.

### 3 Supporting Text S3: Reopening strategies in detail

In this supplementary text we provide a detailed description of each reopening strategy considered.

(i) **Isolation of year group bubbles.** Under this strategy, upon the identification of a case (through a symptomatic pupil seeking a PCR test), all pupils within a year-group bubble were placed in isolation for ten days following the last contact with the positive case. This approach was widely adopted by schools in the UK from September to December 2020. By doing so, any secondary infections from the infected individual would also be isolated, hence halting within-school transmission within the year group. An obvious drawback of such a strategy, however, is the significant number of absences it can cause.

(ii) **Serial contact testing.** Under this strategy, all pupils were tested twice (with LFTs) in the week before returning to school. On the identification of a case (following confirmation by PCR), all pupils within a year-group bubble were tested using LFT for seven days following the last contact with the positive case. The period of serial contact testing is reset if another pupil in the year-group bubble returned a positive LFT result. If the index case pupil tested positive to a confirmatory PCR test, those in the year-group bubble not in isolation were set to undergo a test for the next seven days. If the index case pupil tested negative to the confirmatory PCR test, the period of serial contact testing was only extended until the day the negative test was returned (we assumed a fixed two day delay between date of PCR test and result being known). School testing did not occur at weekends - for serial contact testing days that fell on a weekend, pupils were assumed to be in isolation.

(iii) **Regular mass testing.** As for the serial contact tracing strategy, all pupils were tested twice in the week before returning to school. Once terms begin, pupils were tested once a week, irrespective of whether there had been any confirmed cases within the school. A different year group underwent testing on each day. Any positive cases that were identified took a confirmatory PCR tests with the need to continue in isolation dictated according to the result of that test. However, positive tests did not result in any further school testing, nor in the isolation of year group bubbles.

(iv) **A combined strategy of regular mass testing and serial contact testing implemented in tandem.** Again, pupils were tested twice in the week before returning to school. Identification of positive cases, either through weekly mass tests, or symptomatic pupils seeking a PCR test, triggered the onset of serial contact testing as in (ii).

(v) **No school-level testing or isolation of year-group bubbles.** Finally, we considered a situation in which a school implemented a control strategy involving no testing, and no isolation of year group bubbles. Other than symptomatic individuals isolating when seeking a PCR test, and isolating for 10 days from the test upon receiving a positive test, no testing measures to control the spread of within-school infection were included. We considered this situation to enable assessment of the effectiveness of other strategies at reducing within-school transmission.

### 4 Supporting Text S4: Sensitivity analyses

There is considerable uncertainty surrounding many of the parametric assumptions that underpin the model. Accordingly, we performed a univariate sensitivity analysis to understand the impact these assumptions have on our findings (Fig. S2, Fig. S3, Fig. S4).

In these sensitivity analyses, we compared the relative impact of strategies involving rapid testing to a strategy of isolating year group bubbles. Specifically, we considered: the percentage of simulations in which there were more cases under the rapid testing strategy than under an isolation of year group bubbles strategy; the relative increase in infection compared to an isolation of year group bubbles strategy; the percentage of asymptomatic cases identified through rapid testing for each strategy; and the relative reduction in absences compared to an isolation of year group bubbles strategy.

We performed the sensitivity analysis using a one-at-a-time procedure, with one parameter varied from its baseline value whilst all other parameters were kept constant. We defined a lower than baseline parameterisation and higher than baseline parameterisation for each factor considered. Where parameters were drawn from a distribution for the main analysis, the midpoint value of that parameter was taken as the baseline value for the sensitivity analysis (Table 1).

Across the range of alternative parameterisations, serial contact testing (Fig. S2) and regular mass testing strategies (Fig. S3) remained less effective at reducing infections than an isolation of year group bubbles strategy. Combined regular testing and serial contact testing (Fig. S4) remained more effective at reducing infections in all but one sensitivity scenario considered - for low sensitivity LFTs, the impact of combined testing was approximately equal to that of an isolation of year group bubbles strategy. Of all the factors considered, the sensitivity of LFTs had the largest impact on infections.

Rapid testing strategies reduced the average number of school days missed per pupil by over 90% across the range of alternative parameterisations considered. Community infections, captured by the external force of infection on individuals, and the level of within-school infection, had the largest impact on the reduction of school days missed. Both increase the total level of infection (unlike Fig. S5, we do not adjust the other parameter to keep the total number of infections under an isolation strategy constant). At higher total levels of infections, the reduction in school days missed is smaller, as a result of the increased number of infections under a serial contact testing strategy.

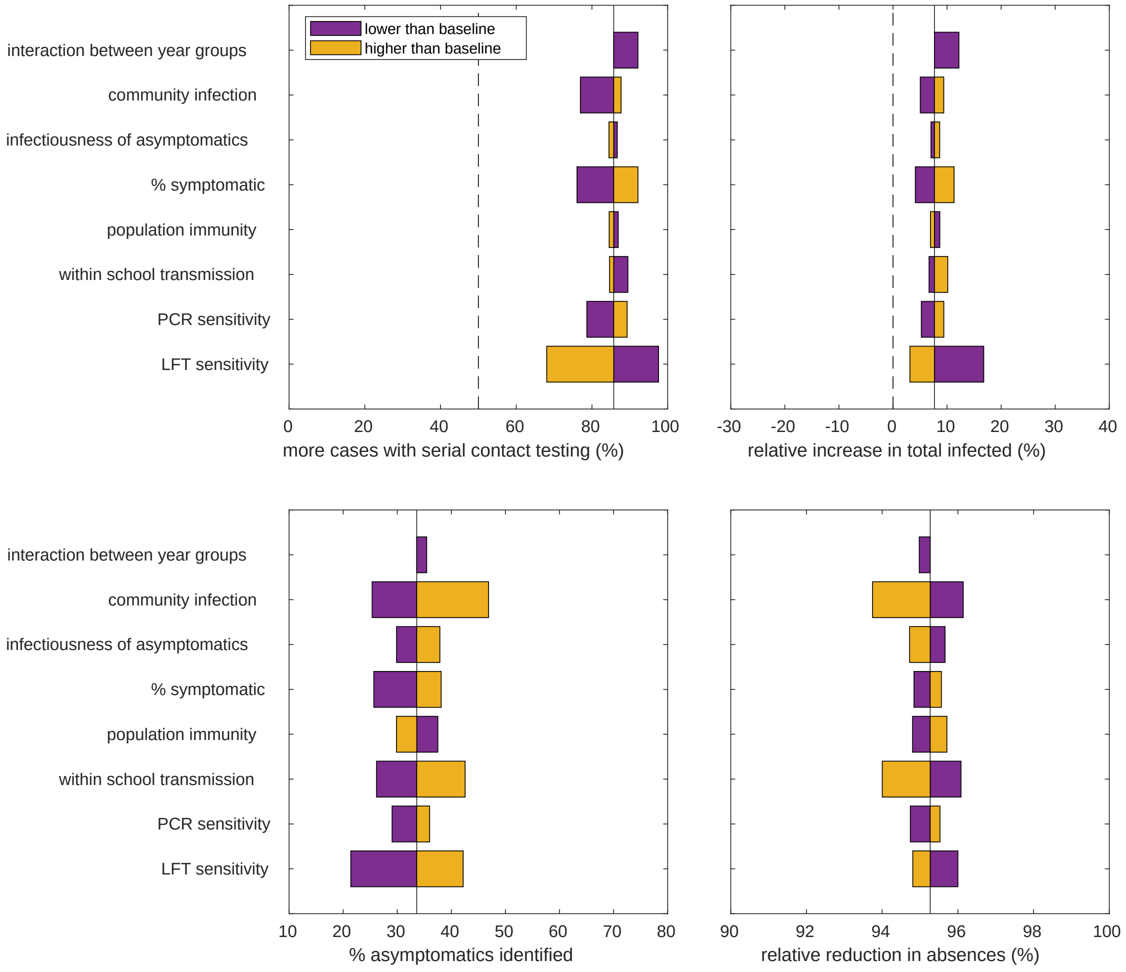


**Fig. S2: Univariate sensitivity analysis of the impact of serial contact testing compared to isolation of year group bubbles.** The tornado diagrams above show univariate sensitivity analyses on the impact of a serial contact testing strategy in comparison to a strategy of isolating year group bubbles. Specifically, we consider: **(top left)** the percentage of simulations with more cases under a mass testing strategy compared to a strategy of isolating year group bubbles; **(top right)** the expected increase in cases over a half term; **(bottom left)** the percentage of infected asymptomatic individuals identified through rapid testing; **(bottom right)** the reduction in school days missed from a policy in mass testing. Under all sensitivity scenarios considered, a serial contact testing strategy was found to be less effective at controlling the spread of infections within the school community than an isolation strategy, did not capture the majority of asymptomatic individuals, but was consistently effective at reducing the number of school days missed. Out of all the factors considered, LFT sensitivity had the largest impact on infection levels.

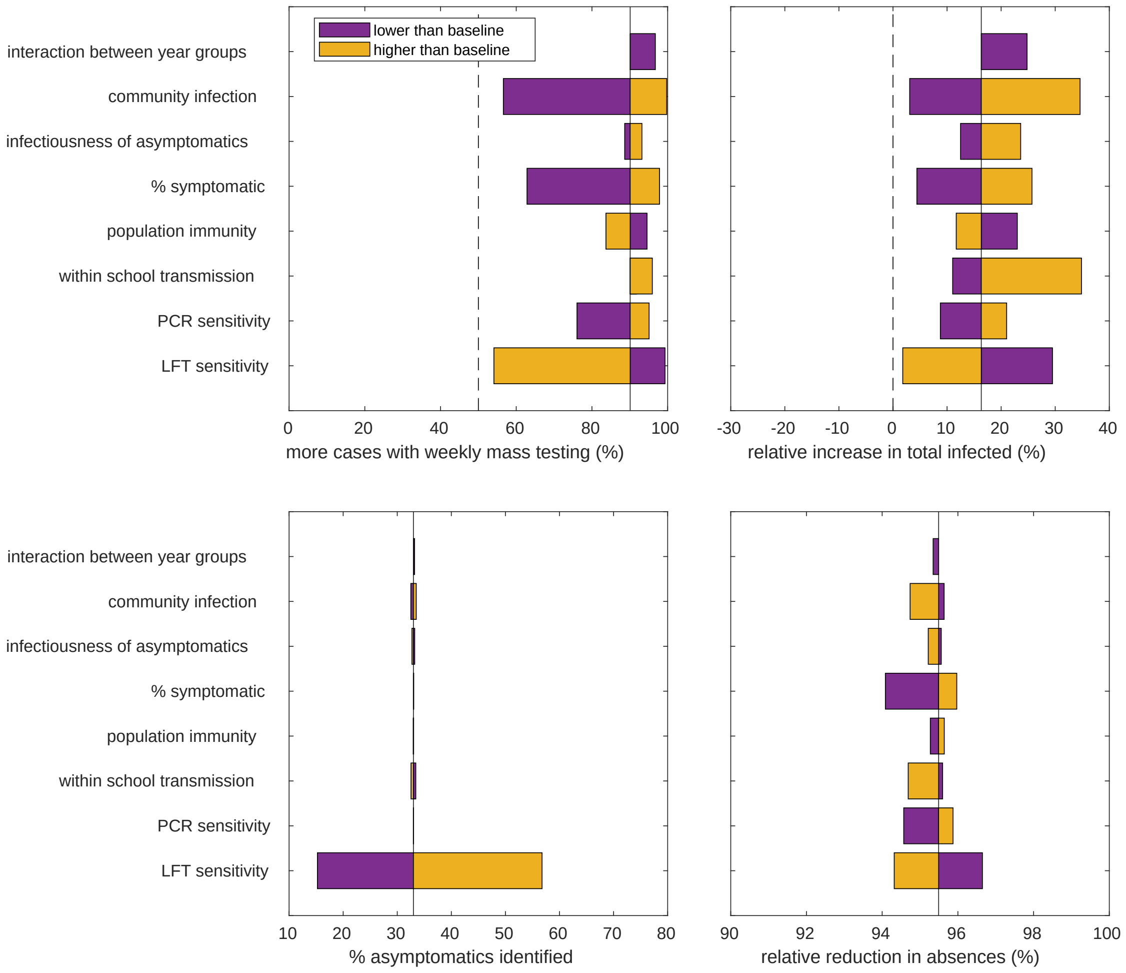


**Fig. S3: Univariate sensitivity analysis of the impact of weekly mass testing compared to isolation of year group bubbles.** The tornado diagrams above show univariate sensitivity analyses on the impact of a weekly mass testing testing strategy in comparison to a strategy of isolating year group bubbles. Specifically, we consider: **(top left)** the percentage of simulations with more cases under a mass testing strategy compared to a strategy of isolating year group bubbles; **(top right)** the expected increase in cases over a half term; **(bottom left)** the percentage of infected asymptomatic individuals identified through mass testing; **(bottom right)** the reduction in school days missed from a policy in mass testing. Under all sensitivity scenarios considered, a mass testing strategy was found to be less effective at controlling the spread of infections within the school community than an isolation strategy, did not capture the majority of asymptomatic individuals, but was consistently effective at reducing the number of school days missed.

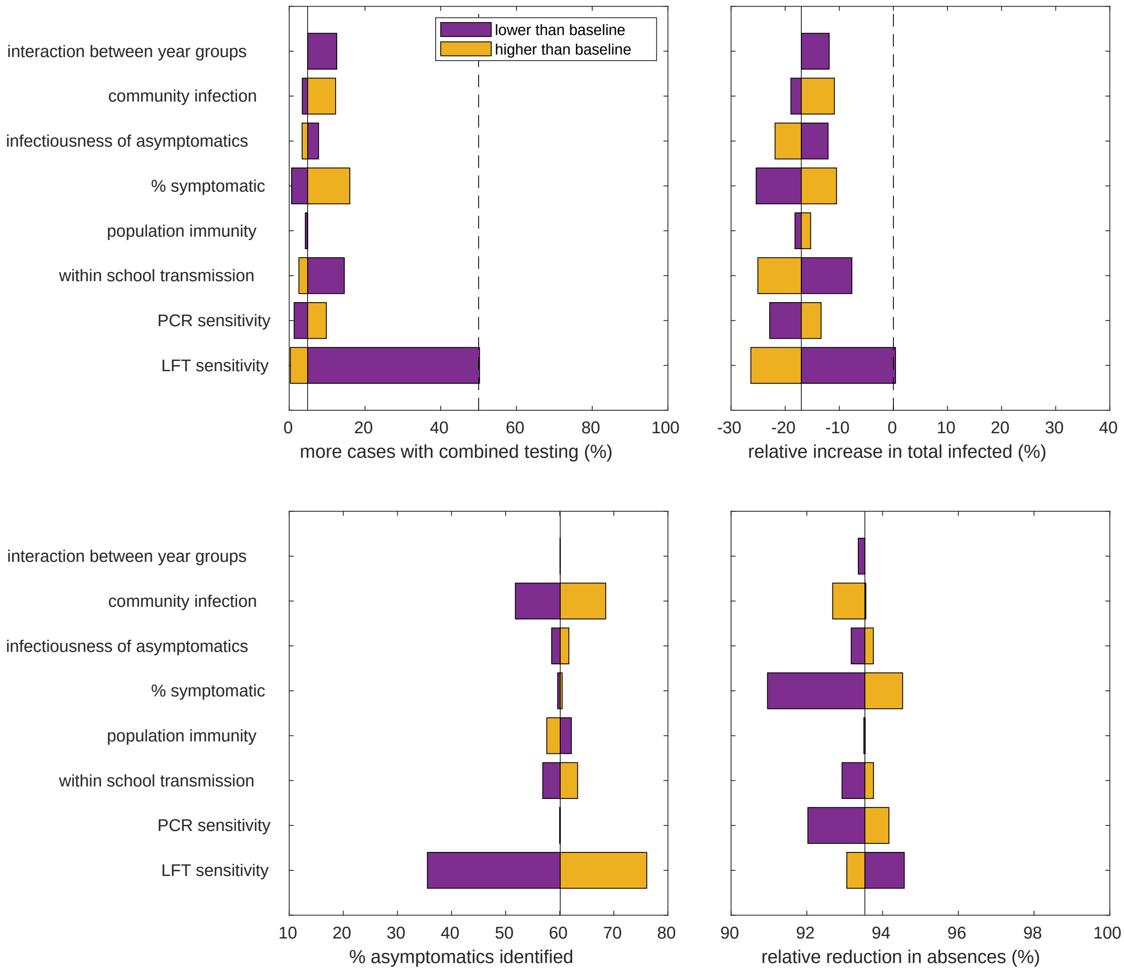


**Fig. S4: Univariate sensitivity analysis of the impact of combined weekly mass testing and serial contact testing compared to isolation of year group bubbles.** The tornado diagrams above show univariate sensitivity analyses on the impact of a combined weekly mass testing and serial contact testing strategy in comparison to a strategy of isolating year group bubbles. Specifically, we consider: **(top left)** the percentage of simulations with more cases under a mass testing strategy compared to a strategy of isolating year group bubbles; **(top right)** the expected increase in cases over a half term; **(bottom left)** the percentage of infected asymptomatic individuals identified through mass testing; **(bottom right)** the reduction in school days missed from a policy in mass testing. In all sensitivity scenarios considered but one, a combined weekly mass testing and serial contact testing strategy was found to be more effective at controlling the spread of infections within the school community than an isolation strategy, captured the majority of asymptomatic individuals. This strategy was only less effective than isolation for low LFT sensitivity, where epidemiological outcomes are comparable. This strategy was consistently effective at reducing the number of school days missed.

### 5 Supporting Text S5: Level of within-school transmission

The levels of transmission within a school setting remains uncertain and will be influenced by a number of factors, including the transmissibility of circulating strains and the efficacy of within-school distancing measures. We considered the impact that reopening strategies had at a range of values of $K$, the parameter determining within-school transmission. To understand the impact of within-school transmission specifically, we kept the total level of infection constant. For a given value of $K$, we adjusted the external source of infection, $\epsilon$, so that an average of 10% of pupils had been infected over a half-term under an isolation strategy.

At the extremes of values considered, the infection dynamics through time stand in contrast with one another. At $K = 0$ (Fig. S5(a)), no transmission occurs within the school. At this value, the only benefit conferred through any strategy is through the isolation of individuals removing their external probability of infection. As a consequence, control strategies have relatively little impact on prevalence. At $K = 4$ (Fig. S5(b)), around 70% of infections occur within school under an isolation of year-group bubbles strategy. As a consequence of the high level of transmission within the school, prevalence increases markedly as the term progresses for most strategies, with the early identification and isolation of infected individuals becoming more effective at reducing infections.

Across the range of within-school transmission considered, serial contact testing and regular mass testing strategies alone were less effective at reducing the total number of infections within a school than an isolation of year group bubbles strategy (Fig. S5(c)), caused a higher peak prevalence (Fig. S5(d)), did not capture the majority of asymptomatic infected individuals (Fig. S5(e)), but resulted in a lower amount of days absent per student (Fig. S5(f)).

However, serial contact testing alongside regular mass testing resulted in a lower total number of infections and peak prevalence, with the relative benefit of such a strategy increasing with the level of transmission that occurs within-school (i.e. increasing $K$). This is because, for higher amounts of transmission occurring within-school, the benefits of capturing a high proportion of asymptomatic infections are amplified. Under this policy, absences are substantially reduced in comparison to a strategy of isolating year groups (Fig. S5(f)). However, pupils do still spend a substantial number of days isolating, although under this strategy, days spent isolating are concentrated over weekends.

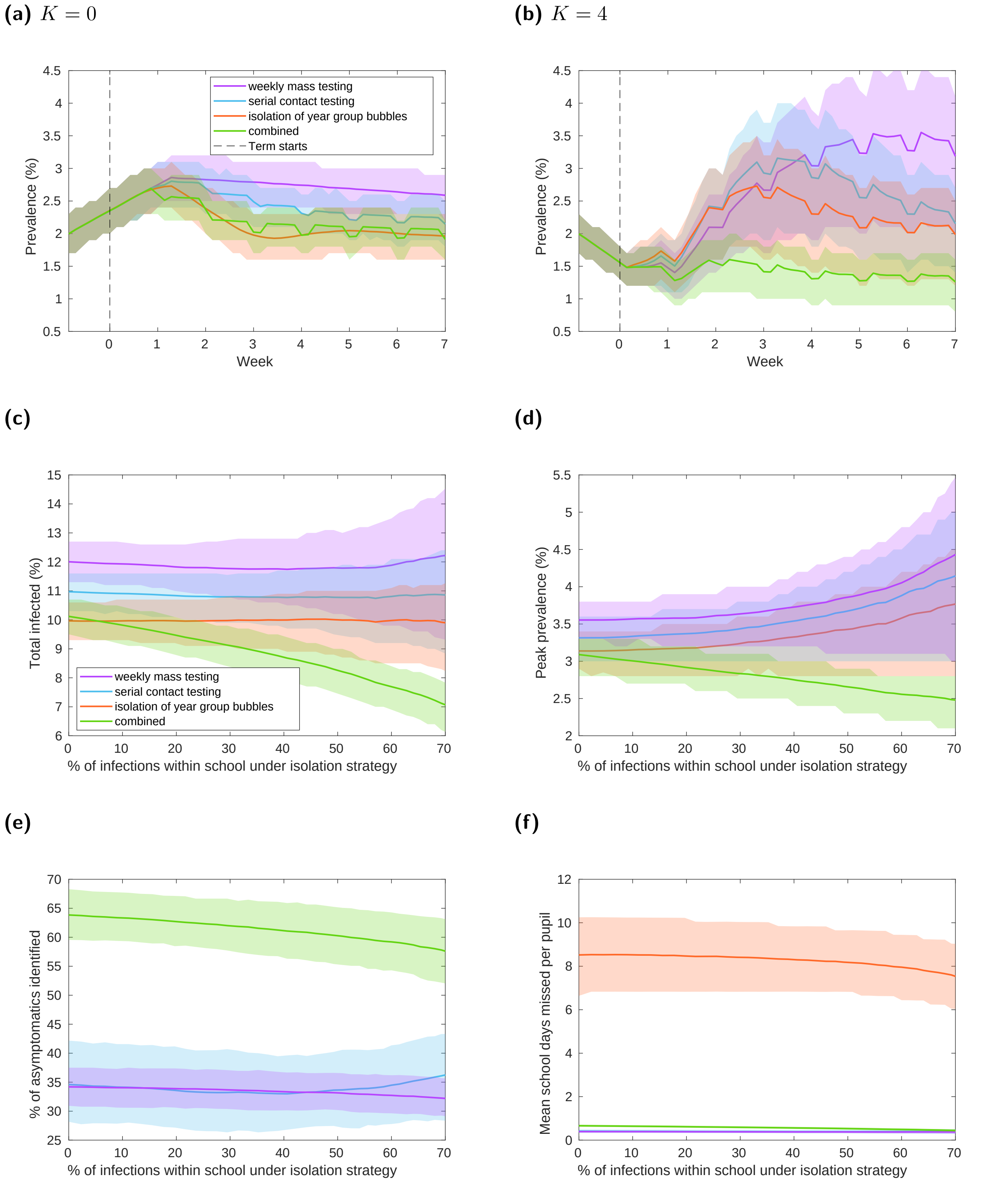


**Fig. S5: Exploring the impact of weighting within-school infection and external infection.** For a given value of $K$, which defines the within-school infectivity, the external force of infection is chosen such that 10% of the school population have been infected by the end of term under an isolation of year groups strategy. In each panel, solid line traces correspond to the mean value attained on each daily timestep and shaded envelopes represent the 50% prediction intervals. The strategies displayed are: weekly mass testing (purple), serial contact testing (blue), year group bubbles strategy (orange), combined serial contact testing and mass testing (green). **(a)** Prevalence given each strategy and $K = 0$. **(b)** Prevalence given each strategy and $K = 4$. **(c)** Percentage of schools pupils infected - a sole mass testing strategy resulted in a larger total number of infections. **(d)** Peak prevalence - on average, a sole mass testing strategy resulted in the greatest peak number of infections. **(e)** Percentage of asymptomatics identified. **(f)** Mean school days missed per student - rapid testing strategies considerably reduces the average number of days students spend isolating.
